# Assessing the Performance of Artificial Intelligence on Anesthesiology In-Training Examinations and Applicability in Medical Education

**DOI:** 10.64898/2026.09.21.26363591

**Authors:** Andrew F. Ibrahim, John F. Zaki

**Affiliations:** School of Medicine, Texas Tech University Health Sciences Center, Lubbock, Texas, USA; Department of Anesthesiology, Critical Care and Pain Medicine, McGovern Medical School at UTHealth Houston, Houston, Texas, USA

**Author notes:** Corresponding Author: Andrew F. Ibrahim, BS, School of Medicine, Texas Tech University Health Sciences Center, Lubbock, TX 79430, USA. **Ethics Statement** This study did not involve human subjects, human samples, or identifiable human data. The analysis used only commercially published question-bank content and machine-generated model responses. Review by an institutional review board or research ethics committee was therefore not applicable to this work, and no approval or exemption determination was sought. No patient or participant data were collected and no informed consent was required. **Data Availability Statement** All model responses, per-item scoring, prompts and analysis scripts are available from the corresponding author on reasonable request. The source question bank is commercially published and is not redistributed.

**Keywords:** artificial intelligence, board examination, graduate medical education, in-training examination, large language models, multiple-choice questions

## Abstract

**INTRODUCTION:** Large language models (LLMs) have demonstrated substantial performance on medical licensing and board examinations, but their application to anesthesiology remains less well studied. Prior evaluations have not compared current-generation models from different developers or assessed performance on figure-dependent questions that earlier models lacked the capability to interpret. This study evaluates the performance of two current-generation LLMs on a comprehensive anesthesiology in-training examination (ITE) review question bank, including figure-dependent items.

**METHODS:** A total of 1,001 single-best-answer questions from an anesthesiology ITE review text, spanning 11 content chapters, were administered to Claude Opus 5 and GPT-5.6. No questions were excluded. Each item was presented once in a fresh, stateless context with no tool access, retrieval, answer key, or explanation provided in the prompt. Performance was analyzed by question format, and all 27 figure-dependent items were administered with their published figures supplied. Accuracy between models was compared using McNemar’s test.

**RESULTS:** Claude Opus 5 answered 947/1,001 items correctly (94.6%; 95% CI, 93.0–95.8), and GPT-5.6 answered 946/1,001 correctly (94.5%; 95% CI, 92.9–95.8); the difference was not significant (McNemar p=1.00). Accuracy was similar across most question formats. On standard questions, accuracy was 94.3% and 94.6% for Claude Opus 5 and GPT-5.6, respectively; on the 18 figure-based questions with figures supplied, accuracy was 94.4% and 83.3%, respectively; and both models achieved 100% accuracy on image-option items. Withholding figures from the same 18 figure-based questions reduced pooled accuracy from 88.9% to 52.8% (exact McNemar p=0.03 and p=0.04 for Claude Opus 5 and GPT-5.6, respectively). The models agreed on 957/1,001 items (95.6%). Of the 33 items both models answered incorrectly, they selected the same incorrect option on 32 (97%).

**DISCUSSION:** Current-generation LLMs achieved approximately 95% accuracy, substantially improving on our prior results with earlier-generation models and placing both models within the highest band of the ITE normative framework, corresponding approximately to the 99th percentile across training levels. By comparison, our prior work with previous models placed ChatGPT-3.5 at the 52nd, 3rd, and 1st percentiles and ChatGPT-4.0 at the 99th, 95th, and 84th percentiles across increasing levels of training. Multimodal capability now permits successful interpretation of many figure-dependent questions, although performance declines markedly when required visual information is unavailable, and highly concordant errors remain. These findings support an increasingly promising role for LLMs as accessible, on-demand educational adjuncts for anesthesiology trainees, provided complete source material is supplied and outputs are critically reviewed.

## INTRODUCTION

Large language models (LLMs) are artificial intelligence systems trained on large text corpora to interpret and generate natural language. They have demonstrated strong performance across a range of medical tasks, including diagnostic reasoning and medical licensing examinations.^1–3^

The Anesthesiology In-Training Examination (ITE) is administered annually to residents, fellows and interns to assess knowledge during training, and performance has been associated with subsequent success in certification.^7^ Despite growing interest in LLMs, literature specific to anesthesiology remains limited and has not kept pace with rapid model development. Prior evaluations assessed models from a single developer, were restricted to text-only questions, and excluded image-dependent items.^4,5^ Yet visual information is integral to anesthesiology assessment and practice, including capnograms, pressure–volume loops, electrocardiographic rhythm strips and regional-anesthesia sonoanatomy. Excluding such items therefore leaves an important component of model performance unexamined.

To address these limitations, we evaluated two current-generation LLMs from different developers, Claude Opus 5 (Anthropic, San Francisco, CA, USA) and GPT-5.6 (OpenAI, San Francisco, CA, USA), on a complete anesthesiology board-review question bank. We retained all questions, supplied the published figure for every figure-dependent item, and analyzed performance by question format rather than excluding visual items. As a prespecified manipulation check, the 18 figure-based items were also administered with the figure withheld. We aimed to characterize overall accuracy, performance across content areas and question formats, the effect of figure availability, and inter-model agreement.

## METHODS

We used a preparatory review text for the anesthesiology ITE and the ABA written examinations.^8^ The text is organized into 11 content chapters covering basic science (chapters 1–4) and clinical science (chapters 5–11). Questions were extracted programmatically from the publisher’s digital edition and reconciled one-to-one against the book’s answer key. A total of 1,001 items and 1,001 corresponding answer-key entries were recovered, with no unmatched items in either direction. Two questions appear twice in the text under different item numbers with slightly reworded explanations; both printings were retained and scored as published. No question was excluded from the primary analysis.

Each item was classified into one of four formats: standard single-best-answer; matching, in which several numbered stems share a lettered option list; figure-based, in which the stem explicitly refers to a figure while the answer options are text; and image-option, in which the answer choices are lettered panels within a figure. Classification required an explicit reference to visual material rather than keyword matching, preventing items that merely contained terms such as ‘tracing’ in ordinary prose from being misclassified.

Each item was submitted to both models under the same primary protocol. Prompts contained the stem, answer options and, for matching items, the shared directions; the answer key, the book’s explanation and bibliographic figure credits were removed. The instruction was identical for all items and both models: ‘Please answer the following multiple-choice question correctly and explain the reasoning for your answer’, followed by a required terminal line in the form ‘ANSWER: <letter>‘. Each item ran in a fresh, stateless process with no conversational carry-over, tool access, web search or retrieval, configuration or memory files, and an empty working directory. The system prompt was ‘You are taking a multiple-choice examination’. If a response lacked a parseable answer line, the item was re-requested; transient interface errors were retried and treated as technical failures rather than incorrect responses. Claude Opus 5 was accessed through its first-party command-line interface and GPT-5.6 through an OpenAI-compatible application programming interface endpoint.

For all 27 figure-dependent items—the 18 figure-based and nine image-option items—the corresponding figure was extracted from the source text as the publisher’s raster image and supplied to both models with the question. This constituted the primary analysis. As a prespecified manipulation check, the 18 figure-based items were re-administered with the figure withheld to quantify the effect of missing visual information.

Accuracy is reported as the proportion of items answered correctly with 95% Wilson score confidence intervals. Because both models answered every item, between-model comparisons were fully paired and were performed using the exact McNemar test, both overall and within chapter, content-division and format strata. For the prespecified figure-availability analysis, the figure-supplied and figure-withheld conditions were compared across the same 18 figure-based items using the exact McNemar test separately for each model, with the pooled change reported descriptively. Inter-model agreement is reported as the proportion of items for which both models selected the same option and, among items both answered incorrectly, the proportion for which they selected the same incorrect option. Analyses were performed in Python 3.12, with a two-sided p-value <0.05 considered statistically significant. No adjustment for multiplicity was applied to exploratory chapter-level and format-level comparisons, which are reported descriptively. The study used only published question-bank content and machine-generated responses and did not involve human subjects, human samples or identifiable human data; review by an institutional review board was therefore not applicable.

## RESULTS

Claude Opus 5 answered 947 of 1,001 items correctly (94.6%; 95% CI, 93.0–95.8), and GPT-5.6 answered 946 of 1,001 correctly (94.5%; 95% CI, 92.9–95.8). Of the 43 items answered correctly by only one model, 22 favored Claude Opus 5 and 21 favored GPT-5.6 (exact McNemar p=1.00). These accuracies were higher than the 72.1% previously reported for ChatGPT-4 and 56.2% for ChatGPT-3.5 on a comparable question bank.^4,5^ Accuracy was similar across the two content divisions: 93.4% versus 94.0% in basic science (p=0.81) and 95.3% versus 94.8% in clinical science (p=0.69), for Claude Opus 5 and GPT-5.6, respectively.

Chapter-level accuracy is presented in Table 1 and Figure 1. Performance was high across all chapters, ranging from 88.5% to 98.8% across the 22 model-by-chapter cells, and no chapter-level comparison between models reached statistical significance. The lowest-performing chapters for both models were pharmacology and pharmacokinetics of volatile anesthetics (90.2% and 88.5%), cardiovascular physiology and anesthesia (92.0% and 88.5%), and anesthesia equipment and physics (92.2% for both). The highest-performing chapters included anatomy, regional anesthesia and pain management (97.6% for both) and neurologic physiology and anesthesia (96.8% for both). In the previous ChatGPT-4 benchmark, anesthesia equipment was the lowest-performing chapter at 44.7%; both models in the present study answered 92.2% of items in the corresponding chapter correctly.^5^

**Table 1.** Accuracy of Claude Opus 5 and GPT-5.6 by Content Chapter and Content Division.

| Content chapter | Items (n) | Claude Opus 5 |  | GPT-5.6 |  | p |
| --- | --- | --- | --- | --- | --- | --- |
|  |  | % correct | 95% CI | % correct | 95% CI |  |
| Basic science |  |  |  |  |  |  |
| 1. Anesthesia Equipment and Physics | 90 | 92.2 | (84.8–96.2) | 92.2 | (84.8–96.2) | 1.00 |
| 2. Respiratory Physiology and Critical Care Medicine | 78 | 93.6 | (85.9–97.2) | 96.2 | (89.3–98.7) | 0.63 |
| 3. Pharmacology and Pharmacokinetics of Intravenous Drugs | 152 | 95.4 | (90.8–97.8) | 96.1 | (91.7–98.2) | 1.00 |
| 4. Pharmacology and Pharmacokinetics of Volatile Anesthetics | 61 | 90.2 | (80.2–95.4) | 88.5 | (78.2–94.3) | 1.00 |
| All basic science items | 381 | 93.4 | (90.5–95.5) | 94.0 | (91.1–95.9) | 0.81 |
| Clinical science |  |  |  |  |  |  |
| 5. Blood Products, Transfusion, and Fluid Therapy | 36 | 94.4 | (81.9–98.5) | 94.4 | (81.9–98.5) | 1.00 |
| 6. General Anesthesia | 149 | 95.3 | (90.6–97.7) | 94.6 | (89.8–97.3) | 1.00 |
| 7. Pediatric Physiology and Anesthesia | 76 | 96.1 | (89.0–98.6) | 92.1 | (83.8–96.3) | 0.25 |
| 8. Obstetric Physiology and Anesthesia | 83 | 94.0 | (86.7–97.4) | 98.8 | (93.5–99.8) | 0.13 |
| 9. Neurologic Physiology and Anesthesia | 62 | 96.8 | (89.0–99.1) | 96.8 | (89.0–99.1) | 1.00 |
| 10. Anatomy, Regional Anesthesia, and Pain Management | 127 | 97.6 | (93.3–99.2) | 97.6 | (93.3–99.2) | 1.00 |
| 11. Cardiovascular Physiology and Anesthesia | 87 | 92.0 | (84.3–96.0) | 88.5 | (80.1–93.6) | 0.25 |
| All clinical science items | 620 | 95.3 | (93.4–96.7) | 94.8 | (92.8–96.3) | 0.69 |
| All items | 1001 | 94.6 | (93.0–95.8) | 94.5 | (92.9–95.8) | 1.00 |

CI: confidence interval. Accuracy is the proportion of items answered correctly, with 95% Wilson score confidence intervals. p values are from the exact McNemar test comparing the two models on the paired items within each stratum. Chapters 1–4 constitute the basic science division and chapters 5–11 the clinical science division. All figure-dependent items were administered with the published figure supplied.

**Figure 1.**
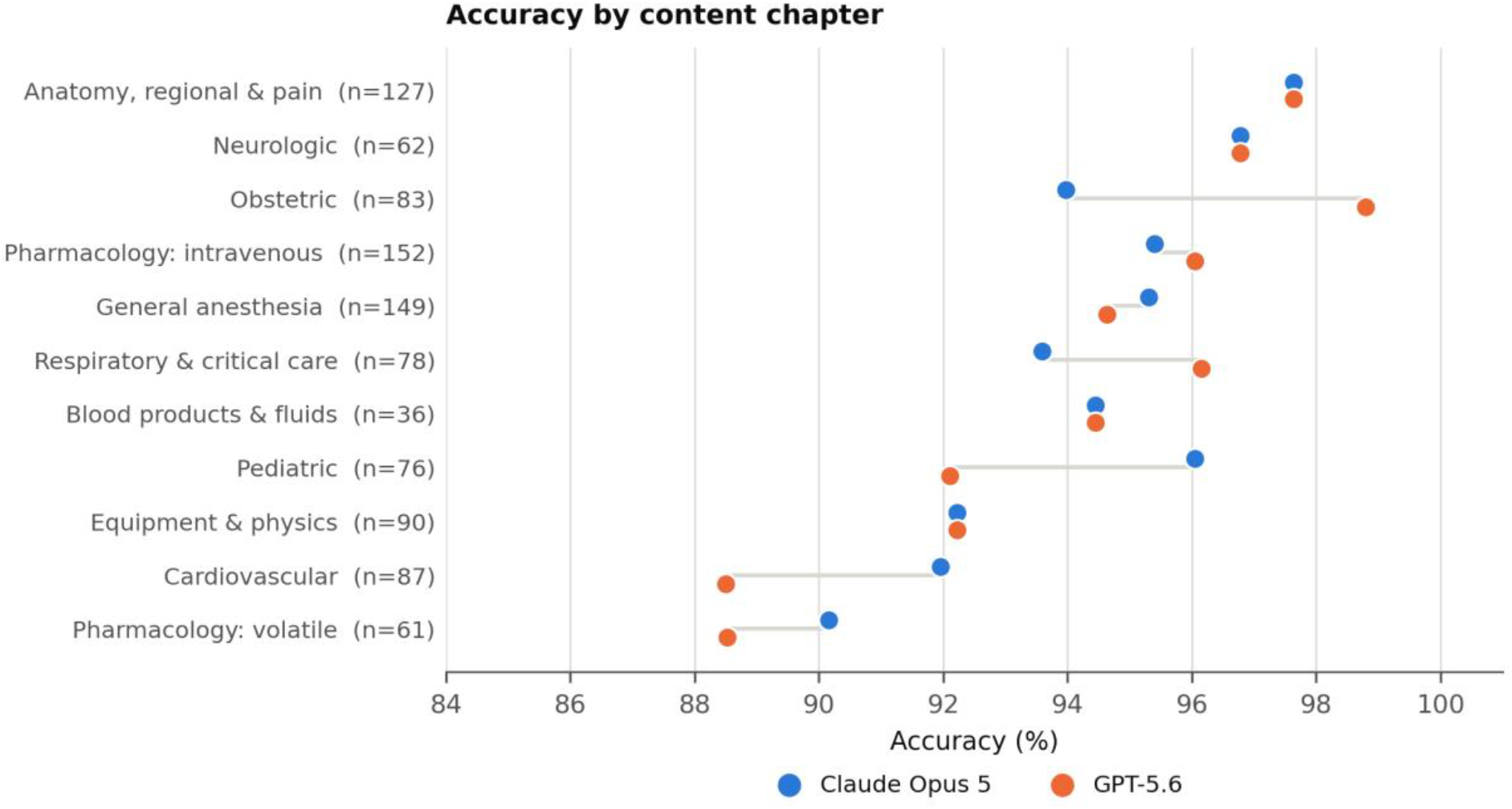
Accuracy by content chapter for both models, ordered by mean accuracy. Chapter denominators are shown in parentheses. No chapter-level comparison between models reached statistical significance.

Accuracy by question format is presented in Table 2. With figures supplied, standard-item accuracy was 94.3% for Claude Opus 5 and 94.6% for GPT-5.6 (n=891); matching-item accuracy was 97.6% and 95.2% (n=83), respectively; figure-based accuracy was 94.4% and 83.3% (n=18), respectively; and both models answered all nine image-option items correctly. Correctly answered figure-dependent items included questions requiring identification of an incompetent expiratory valve from a capnogram, recognition of respiratory variation on an arterial pressure waveform, staging of catheter position using an intravascular electrocardiogram, and identification of individual nerves on axillary sonoanatomy.

**Table 2.**
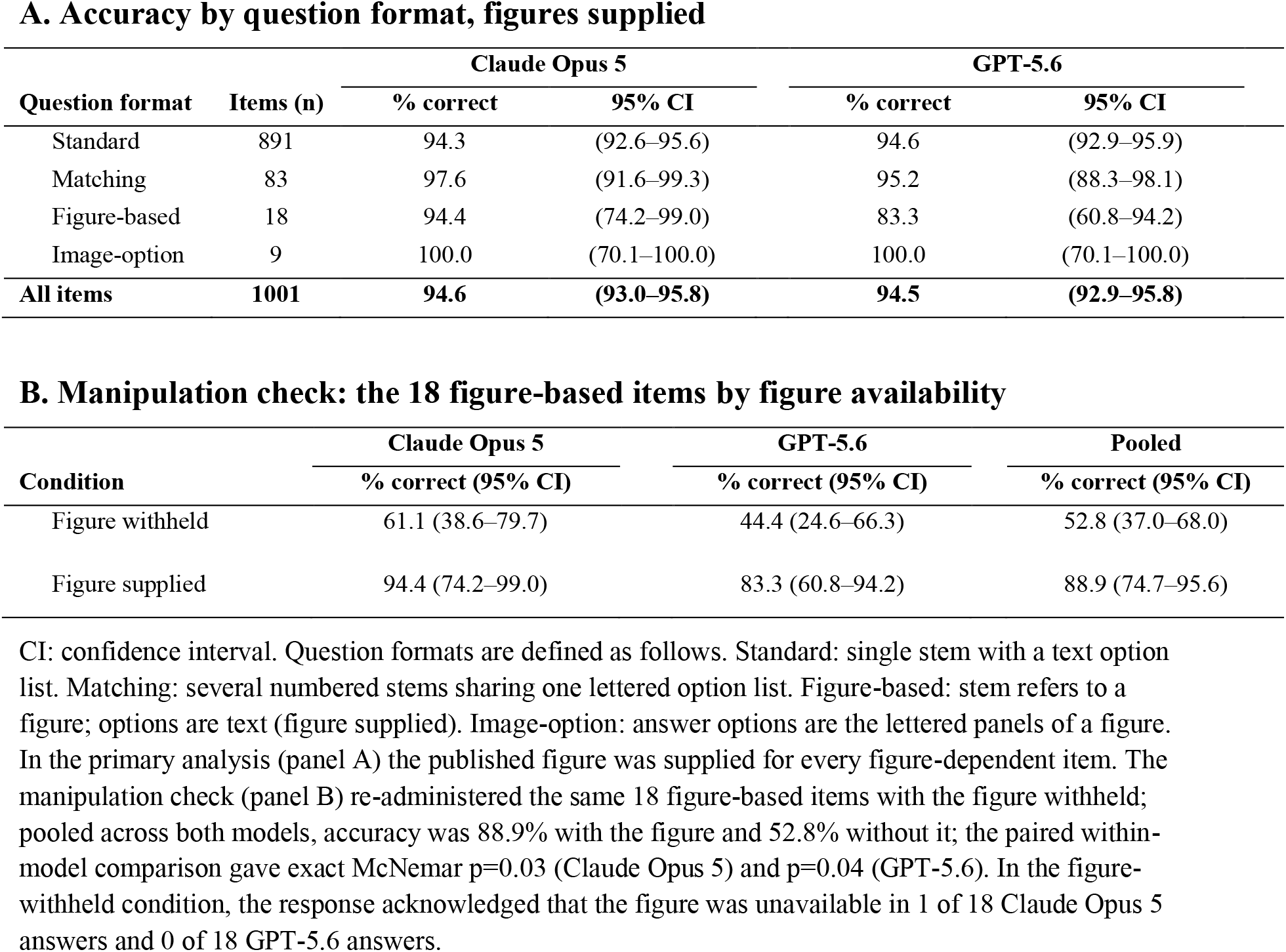
Accuracy by Question Format, and the Effect of Figure Availability on the 18 Figure-Based Items.

**A. Accuracy by question format, figures supplied**
| Question format | Items (n) | Claude Opus 5 |  | GPT-5.6 |  |
| --- | --- | --- | --- | --- | --- |
|  |  | % correct | 95% CI | % correct | 95% CI |
| Standard | 891 | 94.3 | (92.6–95.6) | 94.6 | (92.9–95.9) |
| Matching | 83 | 97.6 | (91.6–99.3) | 95.2 | (88.3–98.1) |
| Figure-based | 18 | 94.4 | (74.2–99.0) | 83.3 | (60.8–94.2) |
| Image-option | 9 | 100.0 | (70.1–100.0) | 100.0 | (70.1–100.0) |
| **All items** | **1001** | **94.6** | **(93.0–95.8)** | **94.5** | **(92.9–95.8)** |

**B. Manipulation check: the 18 figure-based items by figure availability**
| Condition | Claude Opus 5 | GPT-5.6 | Pooled |
| --- | --- | --- | --- |
|  | % correct (95% CI) | % correct (95% CI) | % correct (95% CI) |
| Figure withheld | 61.1 (38.6–79.7) | 44.4 (24.6–66.3) | 52.8 (37.0–68.0) |
| Figure supplied | 94.4 (74.2–99.0) | 83.3 (60.8–94.2) | 88.9 (74.7–95.6) |

CI: confidence interval. Question formats are defined as follows. Standard: single stem with a text option list. Matching: several numbered stems sharing one lettered option list. Figure-based: stem refers to a figure; options are text (figure supplied). Image-option: answer options are the lettered panels of a figure. In the primary analysis (panel A) the published figure was supplied for every figure-dependent item. The manipulation check (panel B) re-administered the same 18 figure-based items with the figure withheld; pooled across both models, accuracy was 88.9% with the figure and 52.8% without it; the paired within-model comparison gave exact McNemar $p=0.03$ (Claude Opus 5) and $p=0.04$ (GPT-5.6). In the figure-withheld condition, the response acknowledged that the figure was unavailable in 1 of 18 Claude Opus 5 answers and 0 of 18 GPT-5.6 answers.

When the same 18 figure-based items were administered without the figure, accuracy decreased from 94.4% to 61.1% for Claude Opus 5 and from 83.3% to 44.4% for GPT-5.6 (pooled 88.9% versus 52.8%, exact McNemar p=0.03 for Claude Opus 5 and p=0.04 for GPT-5.6). This reduction supports a material contribution of the supplied visual information to performance. In the figure-withheld condition, the models rarely acknowledged that the figure was unavailable: explicit acknowledgment occurred in one of 18 Claude Opus 5 responses and none of the 18 GPT-5.6 responses. Both models nevertheless returned an answer and explanation on nearly all figure-withheld items.

The two models selected the same option on 957 of 1,001 items (95.6%). Both answered 925 items correctly and 33 incorrectly. Among the 33 items answered incorrectly by both models, 32 (97%) resulted in selection of the same incorrect option; only one item was answered incorrectly in different directions.

## DISCUSSION

In this evaluation of two current-generation LLMs on a comprehensive anesthesiology board-review question bank organized around the content domains of the ABA written examinations, both models achieved approximately 95% accuracy, with no significant difference in overall performance. Performance remained high across all 11 content chapters, suggesting that these results were broadly distributed rather than driven by a limited number of strong content areas. These accuracies exceed those previously reported for earlier-generation models in anesthesiology.4–6 Notably, anesthesia equipment and physics, previously reported as a relative weakness for ChatGPT-4, reached 92.2% accuracy for both models in the present study.

A principal finding of the present study concerns visually dependent questions. Previous anesthesiology evaluations largely excluded image-based items, leaving an important component of specialty knowledge unexamined.4,5 With the relevant figures supplied, both models performed well on figure-dependent questions, and both correctly answered all nine image-option items. Correctly answered questions required interpretation of capnograms, arterial and central venous pressure waveforms, pressure–volume loops, electrocardiographic tracings, and regional-anesthesia sonoanatomy. These findings suggest that current multimodal LLMs can interpret several forms of visual information encountered in anesthesiology education and extend their potential utility beyond text-only question answering. However, the relatively small number of figure-based and image-option items warrants cautious interpretation of performance within these specific formats.

Figure availability nevertheless had a substantial effect on performance. When the same 18 figure-based questions were presented without their referenced figures, accuracy decreased from 94.4% to 61.1% for Claude Opus 5 (exact McNemar p=0.03) and from 83.3% to 44.4% for GPT-5.6 (p=0.04), corresponding to a pooled decrease from 88.9% to 52.8%. Despite the absence of information explicitly required by the question stem, the models rarely acknowledged that the figure was unavailable and generally still produced a definitive answer accompanied by an explanation. This behavior represents an important practical limitation for educational use. A learner who provides an incomplete question may receive a fluent and seemingly well-reasoned response despite omission of information necessary to answer the question reliably. Referenced visual material should therefore be supplied whenever possible, and responses to figure-dependent questions should be interpreted cautiously when the corresponding image is unavailable. More broadly, educational applications of LLMs may benefit from mechanisms that identify and explicitly flag missing information before generating an answer.

The high degree of inter-model concordance was also notable. Claude Opus 5 and GPT-5.6 selected the same option on 95.6% of all questions, and among the 33 questions answered incorrectly by both models, they selected the same incorrect option on 32. This pattern demonstrates that agreement between independently developed models should not necessarily be interpreted as independent confirmation of correctness. Several explanations are possible, including intrinsically difficult or ambiguous questions, similar learned associations across models, or overlap between benchmark material and model training corpora. Item-level expert review would be necessary to determine whether these shared errors primarily reflect weaknesses in model reasoning, ambiguity in the source material, or other factors.

Several limitations should be considered. First, each question was sampled once under the primary condition, and within-model response stability was not assessed. Some differences between models may therefore reflect stochastic variation rather than stable differences in capability. Second, the study used a commercially published board-review text rather than the ABA ITE itself. Although the material is organized around examination content, the questions are not scaled or equated to the ITE, and raw accuracy cannot be translated directly into an examination percentile or pass–fail determination.

Third, the figure-based and image-option strata contained only 18 and nine questions, respectively, resulting in wide confidence intervals and limiting conclusions regarding relative multimodal performance. Fourth, scoring was based on the final selected answer rather than the factual accuracy, completeness, or pedagogic quality of the accompanying explanation. A correct answer may be supported by incomplete or incorrect reasoning, while an incorrect final choice may follow otherwise useful reasoning. The models also produced substantially different response lengths under the same prompt, further complicating direct comparison of explanation quality. Finally, because the questions were drawn from a commercially published text, prior exposure during model training cannot be excluded. The substantial reduction in performance when figures were withheld argues against simple memorization as a complete explanation for at least some figure-based responses, but it does not eliminate the possibility of training-data contamination.

Viewed alongside our group’s prior work, the improvement across successive generations of LLMs is particularly striking. In our 2024 abstract evaluating 438 anesthesiology ITE review questions, ChatGPT-3.5 achieved 57.6% accuracy and ChatGPT-4.0 achieved 87.2%.9 The present study found accuracies of 94.6% for Claude Opus 5 and 94.5% for GPT-5.6. These findings demonstrate substantial progression in anesthesiology-specific question performance across successive model generations. Our earlier evaluation was also restricted to text-based multiple-choice questions, with image-based assessments excluded, whereas the present study retained figure-dependent questions and demonstrates that current multimodal models can successfully interpret several forms of visual information relevant to anesthesiology.9 Together, these findings suggest that limitations observed in earlier-generation models, including lower overall accuracy and inability to evaluate image-dependent questions, have narrowed considerably with continued model development.

The magnitude of this progression is further illustrated by comparison with resident ITE performance. In our 2024 abstract, model scores were compared with the 2023 ABA ITE percentile norms to provide a clinical training reference. ChatGPT-3.5, with 57.6% accuracy, corresponded to the 52nd percentile for clinical base year residents, the 3rd percentile for second-year residents, and the 1st percentile for third- and fourth-year residents. In contrast, ChatGPT-4.0, with 87.2% accuracy, corresponded to the 99th percentile for clinical base year residents, the 95th percentile for second-year residents, and the 84th percentile for third- and fourth-year residents.9 In the present study, accuracies of 94.6% and 94.5% fall within the highest band of the same normative framework, corresponding to approximately the 99th percentile across levels of training in Supplementary Table 1. Although these comparisons are descriptive and should not be interpreted as direct equivalence to performance on the ABA ITE itself, they provide an intuitive illustration of the scale of improvement observed across model generations.

Taken together, these findings support an increasingly promising role for LLMs in anesthesiology education. Over a relatively short period, model performance on anesthesiology review questions has progressed from substantial variability across generations to consistently high accuracy across content domains, while multimodal capabilities now permit interpretation of visual material that earlier models could not meaningfully evaluate. At the same time, the present study identifies important safeguards for educational use: complete source material should be provided, model agreement should not be assumed to establish correctness, and generated explanations should still be interpreted critically. Future studies should evaluate response reproducibility, explanation quality, calibration and uncertainty expression, performance on prospectively developed questions, and, most importantly, whether LLM-assisted learning improves educational outcomes. With these limitations recognized, current-generation LLMs appear increasingly well positioned to serve as accessible, on-demand educational adjuncts for anesthesiology residents and other trainees.

## Data Availability

Data Availability Statement
All model responses, per-item scoring, prompts and analysis scripts are available from the corresponding author on reasonable request. The source question bank is commercially published and is not redistributed.

## Abbreviations

ABA: American Board of Anesthesiology
AI: artificial intelligence
CI: confidence interval
ITE: In-Training Examination
LLM: large language model.

## SUPPLEMENTARY MATERIAL

Supplementary Table 1. Percentile rank by year of anesthesiology training corresponding to each model’s raw score.

## Declaration of generative AI and AI-assisted technologies in the writing process

The large language models evaluated in this study were the objects of investigation, and their study outputs were generated solely as research data. Generative AI was **not** used in the writing of this manuscript.

**Supplementary Table 1.**
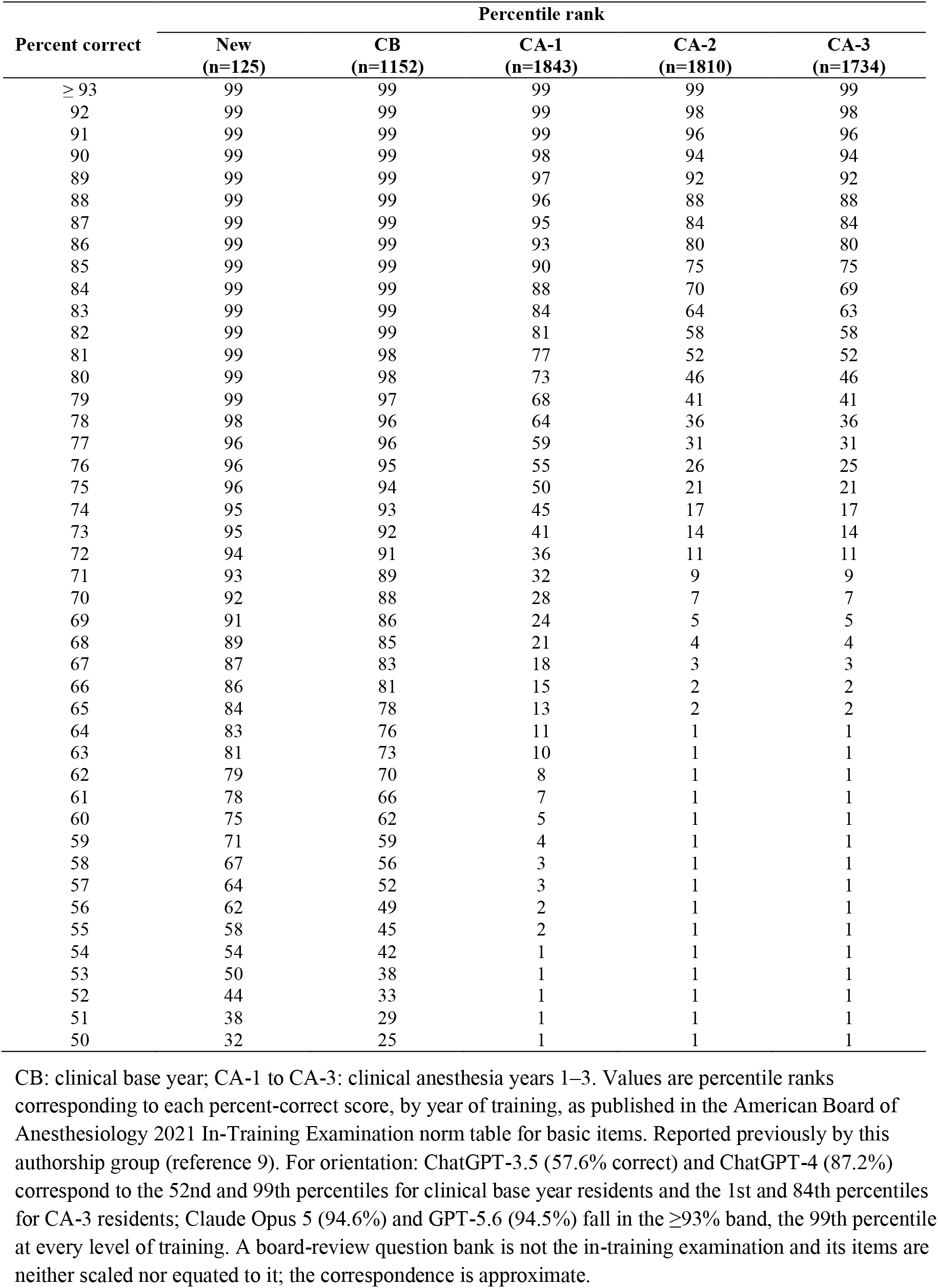
American Board of Anesthesiology 2021 In-Training Examination, Percent Correct Score Norm Table for Basic Items.

