## Supplementary Table 1 for "Assessing the Performance of Artificial Intelligence on Anesthesiology In-Training Examinations and Applicability in Medical Education"

**Supplementary Table 1: American Board of Anesthesiology 2021 In-Training Examination, Percent Correct Score Norm Table for Basic Items**

|  | **Percentile rank** | | | | |
| --- | --- | --- | --- | --- | --- |
| **Percent correct** | **New (n=125)** | **CB (n=1152)** | **CA-1 (n=1843)** | **CA-2 (n=1810)** | **CA-3 (n=1734)** |
| ≥ 93 | 99 | 99 | 99 | 99 | 99 |
| 92 | 99 | 99 | 99 | 98 | 98 |
| 91 | 99 | 99 | 99 | 96 | 96 |
| 90 | 99 | 99 | 98 | 94 | 94 |
| 89 | 99 | 99 | 97 | 92 | 92 |
| 88 | 99 | 99 | 96 | 88 | 88 |
| 87 | 99 | 99 | 95 | 84 | 84 |
| 86 | 99 | 99 | 93 | 80 | 80 |
| 85 | 99 | 99 | 90 | 75 | 75 |
| 84 | 99 | 99 | 88 | 70 | 69 |
| 83 | 99 | 99 | 84 | 64 | 63 |
| 82 | 99 | 99 | 81 | 58 | 58 |
| 81 | 99 | 98 | 77 | 52 | 52 |
| 80 | 99 | 98 | 73 | 46 | 46 |
| 79 | 99 | 97 | 68 | 41 | 41 |
| 78 | 98 | 96 | 64 | 36 | 36 |
| 77 | 96 | 96 | 59 | 31 | 31 |
| 76 | 96 | 95 | 55 | 26 | 25 |
| 75 | 96 | 94 | 50 | 21 | 21 |
| 74 | 95 | 93 | 45 | 17 | 17 |
| 73 | 95 | 92 | 41 | 14 | 14 |
| 72 | 94 | 91 | 36 | 11 | 11 |
| 71 | 93 | 89 | 32 | 9 | 9 |
| 70 | 92 | 88 | 28 | 7 | 7 |
| 69 | 91 | 86 | 24 | 5 | 5 |
| 68 | 89 | 85 | 21 | 4 | 4 |
| 67 | 87 | 83 | 18 | 3 | 3 |
| 66 | 86 | 81 | 15 | 2 | 2 |
| 65 | 84 | 78 | 13 | 2 | 2 |
| 64 | 83 | 76 | 11 | 1 | 1 |
| 63 | 81 | 73 | 10 | 1 | 1 |
| 62 | 79 | 70 | 8 | 1 | 1 |
| 61 | 78 | 66 | 7 | 1 | 1 |
| 60 | 75 | 62 | 5 | 1 | 1 |
| 59 | 71 | 59 | 4 | 1 | 1 |
| 58 | 67 | 56 | 3 | 1 | 1 |
| 57 | 64 | 52 | 3 | 1 | 1 |
| 56 | 62 | 49 | 2 | 1 | 1 |
| 55 | 58 | 45 | 2 | 1 | 1 |
| 54 | 54 | 42 | 1 | 1 | 1 |
| 53 | 50 | 38 | 1 | 1 | 1 |
| 52 | 44 | 33 | 1 | 1 | 1 |
| 51 | 38 | 29 | 1 | 1 | 1 |
| 50 | 32 | 25 | 1 | 1 | 1 |

CB: clinical base year; CA-1 to CA-3: clinical anesthesia years 1–3. Values are percentile ranks corresponding to each percent-correct score, by year of training, as published in the American Board of Anesthesiology 2021 In-Training Examination norm table for basic items. Reported previously by this authorship group (reference 9). For orientation: ChatGPT-3.5 (57.6% correct) and ChatGPT-4 (87.2%) correspond to the 52nd and 99th percentiles for clinical base year residents and the 1st and 84th percentiles for CA-3 residents; Claude Opus 5 (94.6%) and GPT-5.6 (94.5%) fall in the ≥93% band, the 99th percentile at every level of training. A board-review question bank is not the in-training examination and its items are neither scaled nor equated to it; the correspondence is approximate.
